# Smoking Cessation after Transitioning from Hospital to Community Stop Smoking Services: Insights from Real-world Data Analysis

**DOI:** 10.1101/2024.06.05.24308531

**Authors:** Rowan Cherodian, Matthew Franklin, Susan Baxter, James Chilcott, Duncan Gillespie

**Author notes:** Corresponding author: Duncan Gillespie.

## Abstract

**Background:** Patients seen by National Health Service (NHS) hospitals in England should now be asked if they smoke on admission. People who smoke should be treated for tobacco dependence in hospital, then offered support to quit outside of hospital. One way to support post-hospital quitting is through referring patients to community stop smoking services (CSSS). In 2024, the government announced a doubling of CSSS funding for five-years to improve reach and outcomes. Our study aimed to describe the quit rates of people referred from hospital to CSSS, alongside investigating individual characteristics associated with quitting success to inform the potential for more cost-effective, targeted support in the future.

**Methods:** The study was part of a service evaluation using real-world data collected via a CSSS electronic record system in England, which received referrals from hospital-based tobacco teams. We compared CSSS activity and quitting outcomes to local and national reporting data. Generalised Linear Models were used to investigate quitting outcomes 4-weeks after baseline in relation to demographic, socio-economic, nicotine dependence, intervention, and health factors hypothesised to be associated with quitting outcomes.

**Results:** Hospital-referred patients comprised 26% of CSSS referrals, tended to be older, with lower socio-economic status and more long-term health conditions. Overall quitting success by people who made a CSSS supported quit attempt was 61% at 4-weeks, slightly lower than local averages, but similar to national averages. Our analysis sample contained records of 1,326 quit attempts that were supported by CSSS. Of the variables investigated, we found that receiving free NHS prescriptions was consistently associated with lower quitting success (Odds Ratio [OR] 0.55, 95% Confidence Interval [CI] 0.33–0.92), potentially because this is a proxy for lower socio-economic and health status. After accounting for other factors, having cancer relative to no health conditions was associated with higher quitting success (OR 2.26, 95%CI 1.18–4.33).

**Conclusion:** Ensuring patients continue seeking support to quit smoking after their interaction with hospital-based services can lead to quit rates comparable to other CSSS attendees who make a quit attempt. Our analyses highlight the importance of hospital and CSSS investment in improving the transfer of care between services.

## 1. INTRODUCTION

Tobacco smoking remains a leading cause of morbidity and mortality worldwide.^1^ In England, smoking prevalence has been declining since the 1998 “Smoking Kills” White Paper.^2–4^ This decline has been attributed to treating individual tobacco addiction alongside population-level tobacco control measures.^5^ Current government goals are for a smokefree England by 2030, i.e. smoking prevalence <5%.^6^ However, modelling indicates that to reach this <5% goal, more intervention is needed.^5,7^ Interventions should be directed to more disadvantaged groups in which smoking rates remain high, such as those on low-income, unemployed, social housing residents, poverty stricken, and experiencing homelessness.^8,9^

The NHS Long Term Plan aims to offer NHS-funded tobacco dependence treatment services to all identified smokers admitted to NHS hospitals who may not have previously considered quitting smoking.^10^ The logic being that a hospital contact, particularly smoking-related contacts (e.g. cancer diagnosis), can represent an important moment in which people may be more motivated to quit smoking.^11,12^ This service-based intervention involves informing patients identified as smokers about the hospital’s smoke-free policy, then incorporating treatment of their tobacco dependency into their personal care plans.^13,14^ With their consent, they are then offered ongoing support to stop smoking outside of hospital. Evidence suggests that an attempt to quit smoking initiated through a hospital-delivered intervention is more successful when followed-up with at least one month of support.^15^ For example, the Ottawa model for smoking cessation suggests inpatients should receive eight phone calls from the hospital-based tobacco teams over 6-months after discharge^16,17^; the CURE model suggests up to 12-weeks of follow-up support.^18^

However, in England the available support outside hospital varies depending on local resources and policy.^13^ The main option for onwards support are community stop smoking services (CSSS), which since 2013 have been funded by local governments in England and are generally commissioned to support anybody in the local area who is seeking quit support.^19–21^ In 2023, 63% of local governments in England commissioned CSSSs.^22^ With steadily declining investment over the last decade, in 2024 the government committed an additional £350-million over 5-years (£70-million per year) through the “Stopping the start” programme, which would more than double existing funding.^23^ This additional funding is intended to support local governments to improve CSSS reach, accessibility, and outcomes. With new in-hospital tobacco dependence treatment services referring patients to CSSS in certain areas, this new funding will provide an opportunity to explore how CSSS support can be customised to better engage hospital-referred clients, e.g. examining the timing, location and methods through which CSSS support is provided.^19,24^

The current study is part of an evaluation of the QUIT hospital-based tobacco dependence treatment service in South Yorkshire, England (https://sybics-quit.co.uk; Supplementary Information).^25^ The evaluation described in this article focuses on quitting outcomes for people engaged with the QUIT service that are referred to a CSSS and subsequently go on to make a CSSS supported quit attempt,^25^ for which there were three objectives: (1) describe the workload (number of people) impact of the QUIT service on the CSSS; (2) compare quit rates between hospital-referred individuals and the general population using CSSS support; (3) investigate the factors that influenced quitting outcomes among hospital-referred individuals. For (3), we used regression-based analyses to identify factors that might be associated with quitting smoking success, to identify patient or service characteristics that could be targeted to improve quitting outcomes.

## 2. METHODS

### 2.1. The QUIT service and associated study sample

The QUIT service began May 2021 to deliver hospital-based tobacco dependence treatment across four acute hospitals, three specialist mental health, and one children’s hospital, which also includes treatment of tobacco dependency in hospital-based NHS staff, with staggered deployment across hospital sites up to May 2022.^25^ The scope of the service excludes patients on maternity wards who receive support to stop smoking via a different service pathway. Our evaluation data corresponds to patients who had their tobacco dependence treatment care transferred from an acute hospital to the NHS Yorkshire Smokefree CSSS (https://yorkshiresmokefree.nhs.uk/) over a 21-month period: 1^st^ July 2021 to 31^st^ March 2023. The three acute hospitals referring patients to this CSSS were Barnsley Hospital NHS Foundation Trust, Doncaster and Bassetlaw Hospitals NHS Foundation Trust, and Sheffield Teaching Hospitals NHS Foundation Trust.

During the first year of the QUIT service, tobacco treatment advisors were gradually becoming active on hospital wards and the tobacco treatment advisor electronic record systems were beginning to be used.^25^ Subsequently, the QUIT service underwent a series of improvement processes, part of which focused on improving the transfer of patient care for tobacco dependency to CSSS. These patients could have been inpatients or outpatients who had contact with the QUIT hospital-based tobacco team and, if they consented, were referred to CSSS for continued support. Patients could be referred to CSSS after either receiving Very Brief Advice or having a specialist assessment involving motivational interviewing by a hospital-based tobacco dependence treatment advisor.^13^ After discharge, the hospital team routinely calls patients who have had an assessment, confirming the CSSS referral. If a patient identified as a smoker is discharged before seeing the hospital’s tobacco team, they are contacted by phone and offered a specialist assessment and referral to CSSS.

Our evaluation used CSSS real-world data collected for the purposes of service monitoring and improvement, i.e. the CSSS added data fields to their electronic record system that notify them of a hospital-referred client alongside other relevant information (e.g. smoking-related details). The CSSS attempted to contact each referred patient, some of whom were not contactable, and of those who could be contacted, some did not accept the offer of support. Those who did accept the offer of support were registered with the service. However, not everyone who registered went on to make a CSSS support quit attempt. This statistical analysis of quitting outcomes in this study was conducted on the subset of people who made a CSSS supported quit attempt.

### 2.2. Ethical approval

Ethical approval was obtained from the Division of Population Health ethics committee, School of Medicine and Population Health, University of Sheffield (ref: 056472).

### 2.3. Outcome measure: 4-week quit

A “4-week quit” is defined according to the nationally adopted standard, as an individual not having smoked at all in the last 2-weeks when asked at 4-weeks (28-days) from their “quit date”, marking the start of their quit attempt.^26,27^ A person is counted as having achieved a self-reported 4-week quit if they are assessed (face-to-face or by telephone) and state that they have not smoked according to this standard. A person is counted as having achieved a Carbon Monoxide (CO) verified 4-week quit if they self-report having quit and their expired air CO is assessed 4-weeks after their quit date (−3 or +14 days) and found to be less than 10 parts per million. During the study period, CO monitoring of quits was not required due to COVID-19 related restrictions. CO monitoring might also not be appropriate for people feeling unwell, as is likely in our hospital-based sample. Someone was therefore counted as having quit if their quit was either self-reported or CO validated.

For inpatients who had a specialist assessment by a hospital-based tobacco dependence treatment advisor, the hospital discharge date is initially set as the patient’s quit date. If the patient was transferred to the CSSS, and they had smoked since discharge, then a new quit date is agreed with the CSSS. If a patient relapsed back to smoking while under CSSS care, then they could reset their quit date. We used data on quitting outcomes at 4-weeks after setting the latest quit date. Individuals are recorded as either “Quit”, “Lost to Follow-up” (LTF) or “Not Quit”. For our primary analysis all “LTF” were set as “Not Quit”; the influence of this assumption on our results was tested by repeating the analysis with “LTF” people excluded from the analysis sample.

### 2.4. Workload impact on community stop smoking services

To address study objective (1), the NHS Yorkshire Smokefree CSSS provided summary statistics on the numbers of people referred to and engaging with their service, in total and from the QUIT service from July 2021 to March 2023 (Supplementary Information). These data showed the number of people contactable after referral, accepting the service and hence registered, setting a quit date, and achieving a 4-week quit. There were also monthly statistics on referrals, quit dates set, and 4-week quits.

### 2.5. General cohort quitting with community stop smoking services support

To address study objective (2), we compared the age, sex (male/female), and occupation profile of people who made a quit attempt in our analysis sample to national and local profiles using publicly available, national CSSS reporting data from 1^st^ April 2022 to 31^st^ March 2023 (Supplementary Information).^28^ We expect that local CSSS supported 4-week quit rates will be higher than the national average because the Yorkshire and the Humber region (that includes Barnsley, Doncaster and Sheffield) has the highest quit rates nationally, potentially due to high-levels of local CSSS investment.^29^

### 2.6. Explanatory variables for regression analyses

The explanatory variables for regression analysis to address study objective (3) were determined through a systematic process.^25^ First, we conducted a literature review that was designed to identify factors that could be related to quitting outcomes.^30^ Second, we cross-referenced the literature review’s suggested variables with the available CSSS data fields, which we grouped into four categories: “demographic and socio-economic factors”, “tobacco smoking factors”, “intervention characteristics” and “health and healthcare setting”. These categories are described below, with further details in the Supplementary Information.

#### Demographic and socio-economic factors

We investigated the association of age (18–34, 35–44, 45–59, 60+), sex, and occupation on 4-week quits. We also investigated the association with being exempt from NHS prescription payments, given that in England someone is eligible for free prescriptions if they meet certain criteria that are potentially proxies for low socio-economic and health status; i.e. being aged 60+, pregnant or a recent mother, having a specified disability or medical condition, or receiving social welfare benefits.^31^

#### Tobacco smoking factors

The Fagerström score of nicotine dependence was dichotomised into a binary variable representing low (0–4) and high (5+) nicotine dependence.^25^

#### Community stop smoking service support

The support provided by CSSS was measured by the number of support sessions attended and an index that we derived to represent the intensity of NRT supplied.^25^ During the study period, Varenicline was not available as a smoking cessation medication.^32^ In their sessions, people could be recorded as receiving up to two different types of NRT, such as patches, sprays, and lozenges in different quantities, with most clients receiving a combination of types over the course of their sessions. Due to high variability in the types and amounts of NRT given, it was not feasible to account for all variations. Thus, an intensity index of NRT given was calculated by dividing the total number of times NRT was given by the number of sessions attended, with a higher value indicating more intensive pharmacotherapy. We dichotomised this index into: people who on average received ≤1 NRT items per session vs. >1 item.

#### Hospital-based intervention

We summarised each patient’s interaction with the hospital-based service into a single variable with four categories:

i. Specialist assessment conducted in-person during the inpatient stay;
ii. Specialist assessment or Very Brief Advice over the phone post-discharge;
iii. Specialist assessment or Very Brief Advice given as an outpatient;
iv. Unknown.

#### Health and healthcare setting variables

Medical conditions were recorded as binary variables of having (or not) any of 16 medical conditions caused or made worse by smoking with no additional details (e.g. sub-condition type, treatment type, time of diagnosis). For analysis, these 16 conditions were grouped to represent: (i) the government’s Major Conditions Strategy as five binary variables for overarching condition categories:^33^ cardiovascular disease, diabetes, mental ill health, cancer, and chronic respiratory condition; (ii) a co-morbidities categorial variable with three categories: 0, 1–2, 3+ conditions.

### 2.7. Data analysis

Data analysis followed the pre-registered analysis plan,^27^ with refinements to suit the limitations of the available data. The mechanism of missingness with respect to quitting outcomes was assessed using two-way t-tests, the results from which informed what missing data method might be suitable, e.g. if the data is plausibly missing at random, using multiple imputation via chained regression equations would be suitable.^34^ Subsequent statistical analysis of the 4-week quit outcome used Generalised Linear Models with logit link functions. The analysis was based on five model structures that sequentially added additional explanatory variables:

- (Model-1) individual demographic and socio-economic characteristics;
- (Model-2) Model-1 with the Fagerström score for nicotine dependence;
- (Model-3) Model-2 with CSSS support (i.e. support sessions and pharmacotherapy);
- (Model-4) Model-3 with hospital-based contact type (i.e. inpatient vs. post-discharge vs. outpatient);
- (Model 5) Model-4 with health co-morbidities.

The model structures were compared based on the Akaike’s Information Criterion (AIC) and Likelihood Ratio Tests of the difference in residual deviance between models. Statistical effect sizes are reported in terms of adjusted odds ratios (ORs). These odds ratios provide a quantification of association strength between the outcome of interest (4-week quits) and the explanatory variables of interest, holding all other variables in the regression model at their respective means. An odds ratio is interpreted relative to a value of 1, whereby 1 suggests there is no difference in odds between groups being compared. An odds ratio <1 indicates a decrease in odds relative to the comparison group, whereas an odds ratio >1 indicates an increase in odds. Thus, if a factor is associated with a decrease in odds of quitting success the odds ratio is below one, and if it is associated with an increase in quitting success the odds ratio is above one. The threshold for statistical significance was set at 0.05 (two-way) to produce 95% confidence intervals. When a 95% confidence interval overlaps one, this indicates that there is more than a 5% probability that the observation of an effect is due to chance. Reporting of statistical uncertainty around the effect estimates adjusted for the multiple imputation of missing data using Rubin’s Rule.^35^

## 3. RESULTS

### 3.1. Patient flows through hospital and community stop smoking services

Of the 3,223 patients referred to the CSSS from the hospital-based service, 72.0% could be contacted by the CSSS, of whom 72.9% went on to register with the CSSS, and of these 78.8% set a quit date, with a 4-week quit success rate of 61.1% (Supplementary Table S3). For the first 10 months of the hospital-based service implementation, there was a steady rise in the percentage of all referrals to the CSSS originating from hospital (Figure 1). After this initial period, from April 2022, the monthly statistics were relatively stable, with the hospital-based service accounting for 26% of all CSSS referrals (191 referrals/month), translating to 19% of quit dates set (81 quit dates/month) and 17% of 4-week quits (49 quits/month).

**Figure 1.**
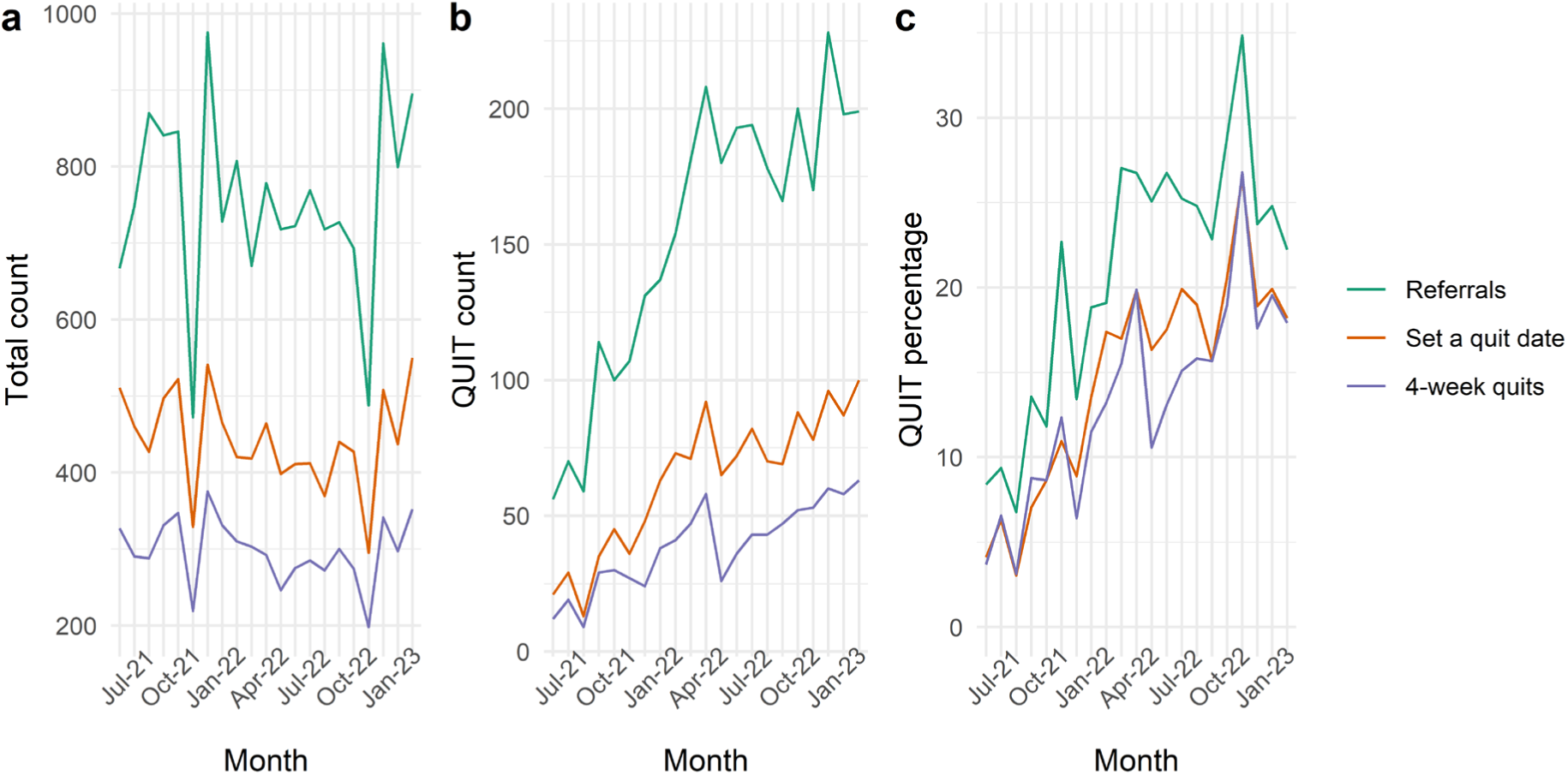
Monthly activity for the community stop smoking service overall and the percentage resulting from hospital referrals.

Put in the context of patient flows through the inpatient tobacco dependence treatment pathway, based on the Key Performance Indicators for the QUIT service from November 2022 to March 2023 (Supplementary Information), for every 100,000 inpatient admissions, there were 9,700 people identified as currently smoking tobacco, of whom 3,700 lived locally and had a specialist assessment by a hospital-based tobacco treatment advisor. Of these specialist assessments, 1,250 people consented to CSSS referral after discharge, resulting in 520 quit dates being set and 320 4-week quits (Supplementary Table S11).

### 3.2. Descriptive statistics and comparison to the national and local samples

Table 1 provides descriptive statistics for the analysis sample with respect to quitting outcomes and the explanatory variables investigated (further details in the Supplementary Information).

**Table 1.** Description of the analysis sample.

|  | Number of observations | Percentage |
| --- | --- | --- |
| **Total** | 1,326 |  |
| **Quitting outcome** |  |  |
| 4-week quit achieved <sup>1</sup> | 813 | 61.3% |
| **Demographic and socio-economic variables** |  |  |
| Sex (male, female) |  |  |
| Male | 691 | 52.1% |
| Age |  |  |
| 18–34 | 112 | 8.4% |
| 35–44 | 190 | 14.3% |
| 45–59 | 504 | 38.0% |
| 60+ | 520 | 39.2% |
| Occupation |  |  |
| Routine and manual occupations | 411 | 31.0% |
| Retired | 321 | 24.2% |
| Sick/disabled and unable to work | 297 | 22.3% |
| Never worked or unemployed for over 1 year | 196 | 14.8% |
| Other | 102 | 7.7% |
| Eligible for free NHS prescriptions | 1,076 | 81.1% |
| **Fagerström score for nicotine dependence (0–4, 5+) <sup>2</sup>** |  |  |
| 0–4 | 542 | 40.8% |
| 5+ | 784 | 59.2% |
| **Community stop smoking service support** |  |  |
| Number of support sessions attended (whole sample average) |  | 6.3 (range: 1 to 26) |
| Nicotine Replacement Therapy items per session (≤1 item, >1 item) |  |  |
| >1 item | 412 | 31.0% |
| **Type of contact with the hospital-based service** |  |  |
| Specialist assessment in-person whilst an inpatient | 584 | 44.0% |
| Post-discharge specialist assessment or Very Brief Advice | 66 | 5.0% |
| Outpatient specialist assessment or Very Brief Advice | 173 | 13.0% |
| Unknown | 503 | 37.9% |
| **Health variables** |  |  |
| Specific health conditions |  |  |
| Chronic respiratory condition | 498 | 37.6% |
| Mental ill health | 349 | 26.3% |
| Cardiovascular disease | 334 | 25.2% |
| Diabetes | 165 | 12.4% |
| Cancer | 123 | 9.3% |
| Number of co-morbid health conditions |  |  |
| 0 | 373 | 28.1% |
| 1–2 | 616 | 46.5% |
| 3+ | 337 | 25.4% |

<sup>1</sup> 10.0% of the sample were lost to follow-up and so had unknown quitting outcomes.
<sup>2</sup> Missing values imputed; statistics generated from the average of 19 imputed data sets, each with $N = 1,326$ .

Compared to the national data on people attempting to quit smoking with CSSS support, our data sample of people referred from hospital included a higher percentage of people who were retired, sick or disabled and unable to work, or eligible for free NHS prescriptions (Supplementary Table S5). Among those arriving from hospital who made a CSSS supported quit attempt, 61.3% achieved a 4-week quit (n=813/1,326; Supplementary Tables S5 and S6). Compared to the local average quit rate with CSSS support of 69.6% (3,540/5,083), the odds of quitting in people who made a quit attempt after being referred from hospital were on average 30.9% lower (χ^2^ =33.1, df = 1, *p* < 0.01; Supplementary Table S6). However, rates of quitting in our data sample were similar to national averages (Supplementary Table S6).

Supplementary Tables S5 and S6 also present these comparisons separately for each demographic and socio-economic subgroup.

### 3.3. Missing data assessment

Across all chosen variables, missing data was mainly associated with the Fagerström score which had 252 missing values (19% of the analysis sample). This missingness was assessed to be “missing at random” with respect to quitting outcomes (non-missing 61.9%, missing 58.7%, *t*-statistic = −1.026, df = 251, *p-*value = 0.306). Missing values for the Fagerström score were therefore imputed using multivariate imputation, based on information from all other analysis dataset variables.^34^ The result was nineteen imputed datasets (same as the percentage missing), accounting for the statistical uncertainty in imputation.

### 3.4. Statistical analyses

Overall, Model-5 with the most complex structure also had the best data fit (Table 2). Table 3 presents the coefficient estimates (odds ratios) for the five model structures considered, with Figure 2 presenting just Model-5’s odds ratios. Model-5’s estimates were robust to assumptions about quitting outcomes for people lost to follow-up and restricting the data sample to only people with no missing Fagerström scores (Supplementary Table S12).

**Figure 2.**
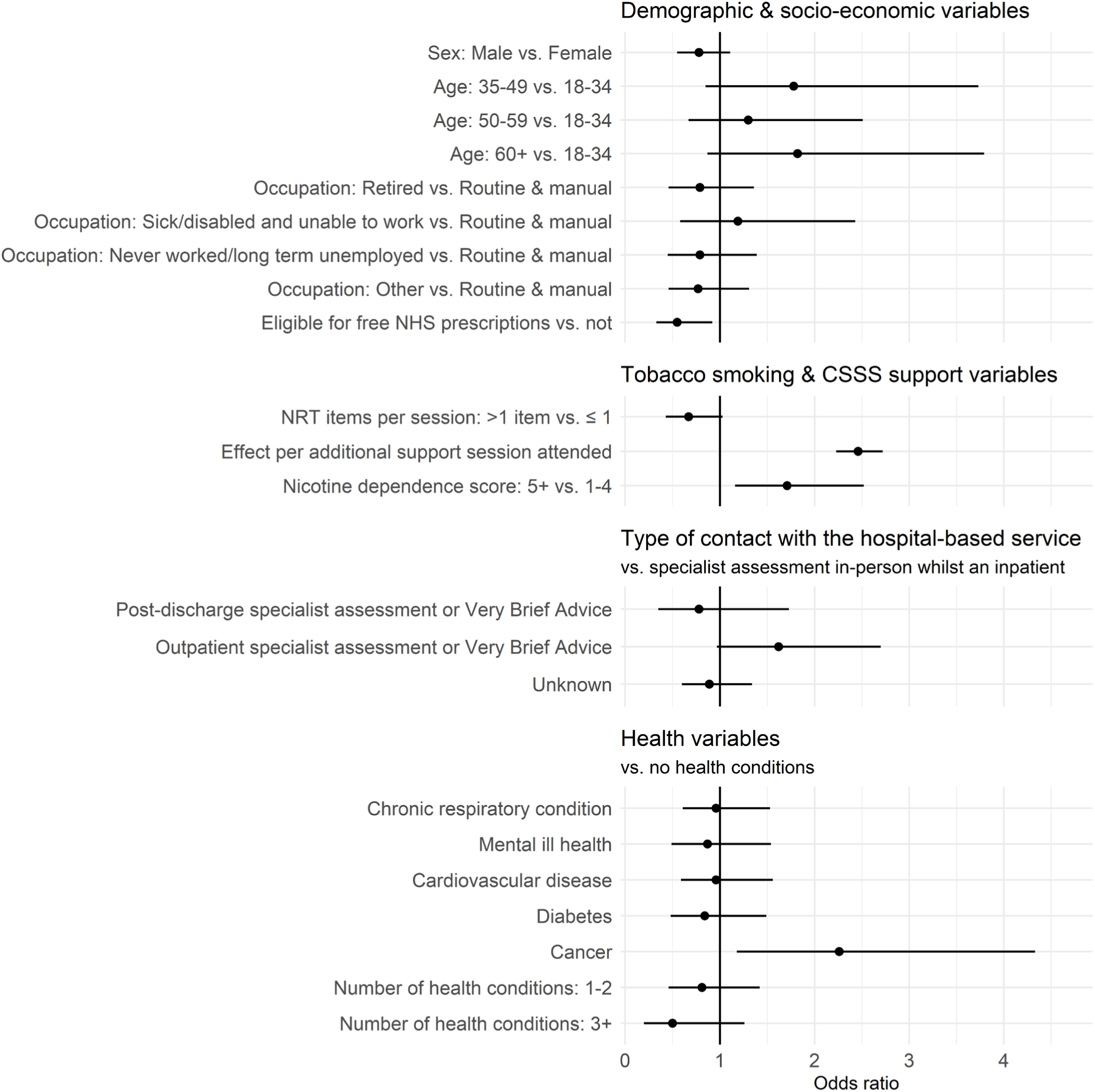
Adjusted odds ratios with 95% confidence intervals showing the relative effects of the explanatory variables from Model 5.

**Table 2.**
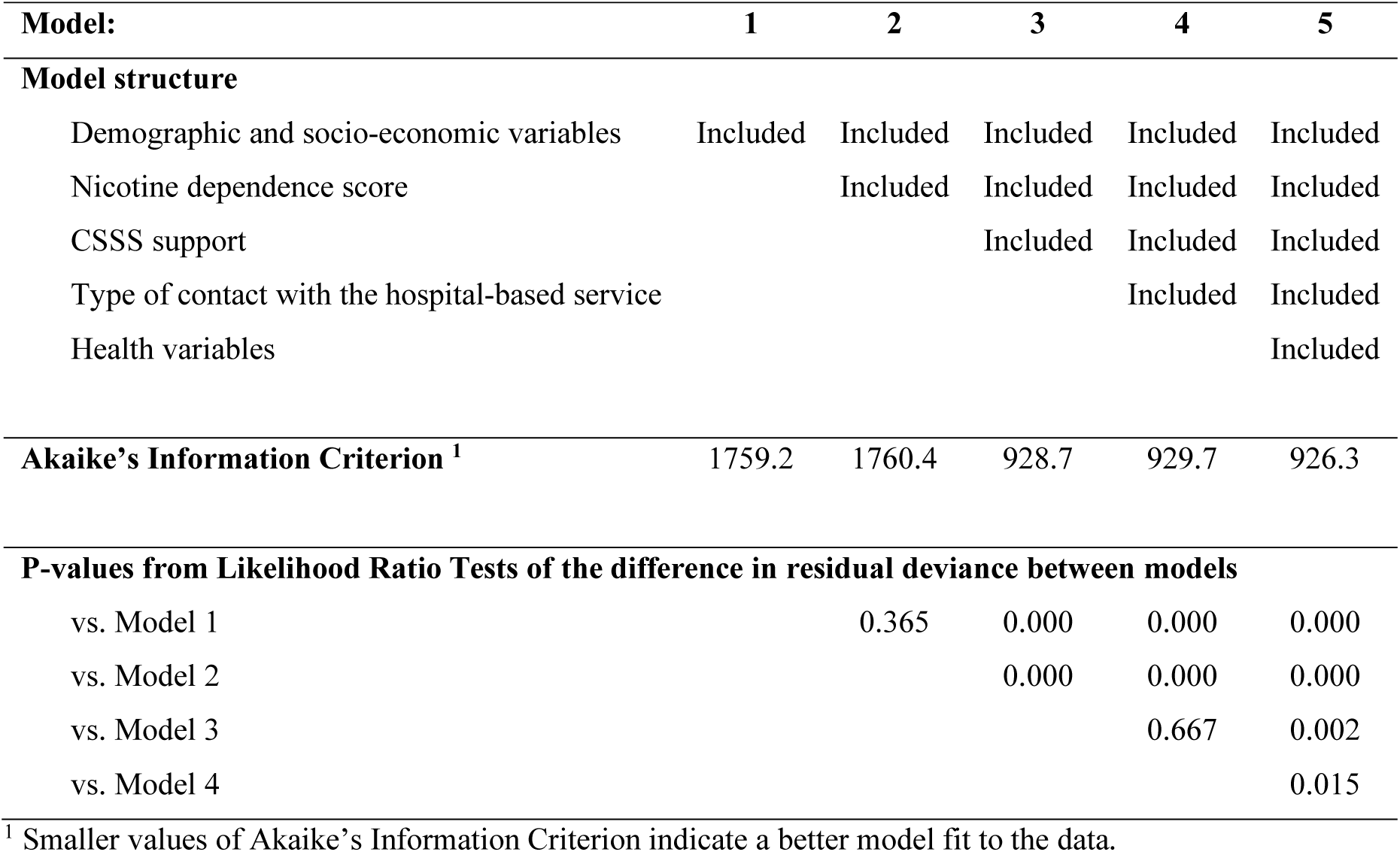
The five alternative model structures investigated and statistics showing comparative model fit.

| **Model:** | **1** | **2** | **3** | **4** | **5** |
| --- | --- | --- | --- | --- | --- |
| **Model structure** |  |  |  |  |  |
| Demographic and socio-economic variables | Included | Included | Included | Included | Included |
| Nicotine dependence score |  | Included | Included | Included | Included |
| CSSS support |  |  | Included | Included | Included |
| Type of contact with the hospital-based service |  |  |  | Included | Included |
| Health variables |  |  |  |  | Included |
| **Akaike's Information Criterion <sup>1</sup>** | 1759.2 | 1760.4 | 928.7 | 929.7 | 926.3 |
| **P-values from Likelihood Ratio Tests of the difference in residual deviance between models** |  |  |  |  |  |
| vs. Model 1 |  | 0.365 | 0.000 | 0.000 | 0.000 |
| vs. Model 2 |  |  | 0.000 | 0.000 | 0.000 |
| vs. Model 3 |  |  |  | 0.667 | 0.002 |
| vs. Model 4 |  |  |  |  | 0.015 |

<sup>1</sup> Smaller values of Akaike's Information Criterion indicate a better model fit to the data.

**Table 3.**
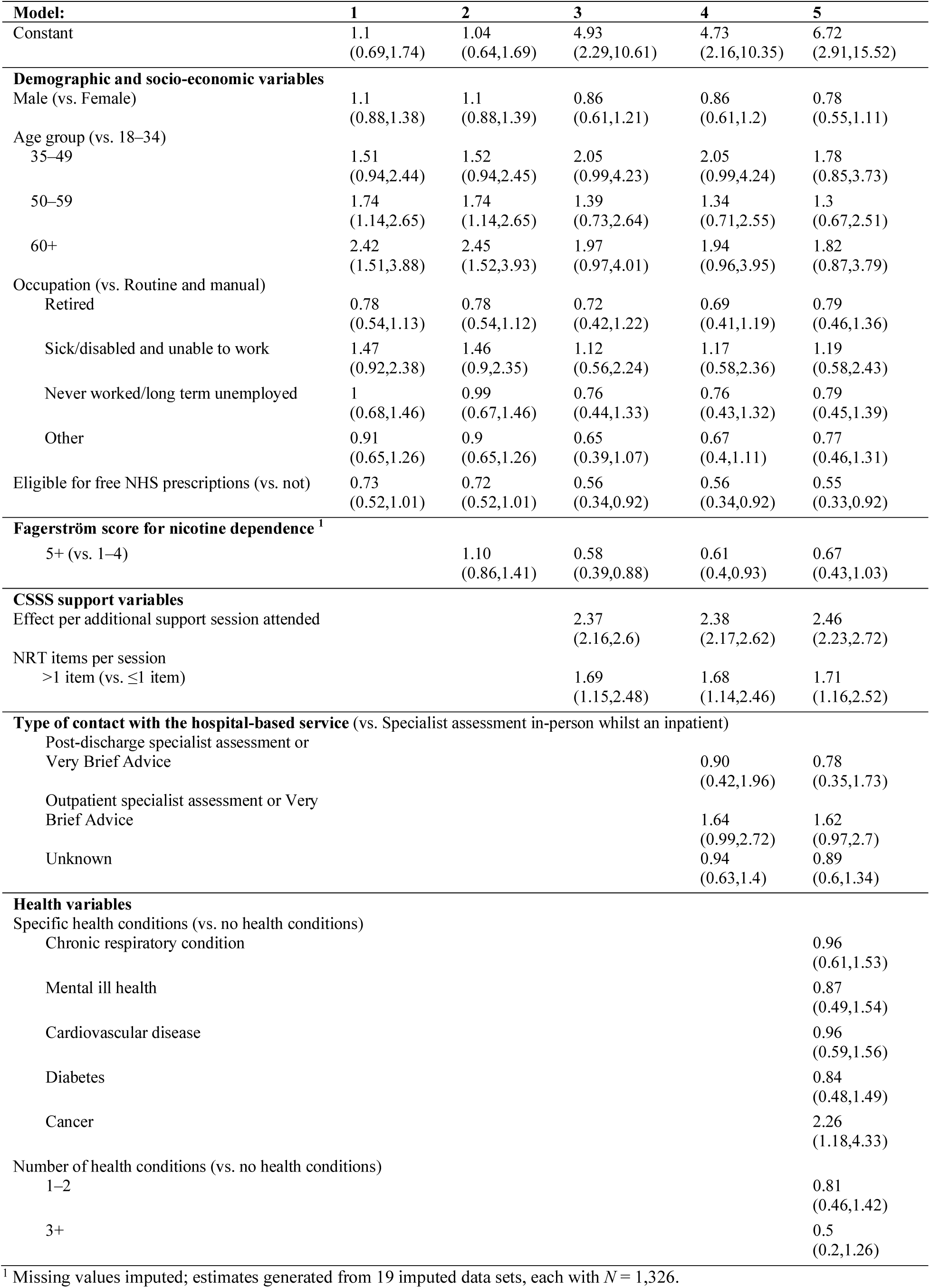
Coefficient estimates (odds ratios) from the five model structures considered. Numbers in parenthesis are 95% confidence intervals.

| **Model:** | **1** | **2** | **3** | **4** | **5** |
| --- | --- | --- | --- | --- | --- |
| Constant | 1.1<br>(0.69,1.74) | 1.04<br>(0.64,1.69) | 4.93<br>(2.29,10.61) | 4.73<br>(2.16,10.35) | 6.72<br>(2.91,15.52) |
| **Demographic and socio-economic variables** |  |  |  |  |  |
| Male (vs. Female) | 1.1<br>(0.88,1.38) | 1.1<br>(0.88,1.39) | 0.86<br>(0.61,1.21) | 0.86<br>(0.61,1.2) | 0.78<br>(0.55,1.11) |
| Age group (vs. 18–34) |  |  |  |  |  |
| 35–49 | 1.51<br>(0.94,2.44) | 1.52<br>(0.94,2.45) | 2.05<br>(0.99,4.23) | 2.05<br>(0.99,4.24) | 1.78<br>(0.85,3.73) |
| 50–59 | 1.74<br>(1.14,2.65) | 1.74<br>(1.14,2.65) | 1.39<br>(0.73,2.64) | 1.34<br>(0.71,2.55) | 1.3<br>(0.67,2.51) |
| 60+ | 2.42<br>(1.51,3.88) | 2.45<br>(1.52,3.93) | 1.97<br>(0.97,4.01) | 1.94<br>(0.96,3.95) | 1.82<br>(0.87,3.79) |
| Occupation (vs. Routine and manual) |  |  |  |  |  |
| Retired | 0.78<br>(0.54,1.13) | 0.78<br>(0.54,1.12) | 0.72<br>(0.42,1.22) | 0.69<br>(0.41,1.19) | 0.79<br>(0.46,1.36) |
| Sick/disabled and unable to work | 1.47<br>(0.92,2.38) | 1.46<br>(0.9,2.35) | 1.12<br>(0.56,2.24) | 1.17<br>(0.58,2.36) | 1.19<br>(0.58,2.43) |
| Never worked/long term unemployed | 1<br>(0.68,1.46) | 0.99<br>(0.67,1.46) | 0.76<br>(0.44,1.33) | 0.76<br>(0.43,1.32) | 0.79<br>(0.45,1.39) |
| Other | 0.91<br>(0.65,1.26) | 0.9<br>(0.65,1.26) | 0.65<br>(0.39,1.07) | 0.67<br>(0.4,1.11) | 0.77<br>(0.46,1.31) |
| Eligible for free NHS prescriptions (vs. not) | 0.73<br>(0.52,1.01) | 0.72<br>(0.52,1.01) | 0.56<br>(0.34,0.92) | 0.56<br>(0.34,0.92) | 0.55<br>(0.33,0.92) |
| **Fagerström score for nicotine dependence <sup>1</sup>** |  |  |  |  |  |
| 5+ (vs. 1–4) |  | 1.10<br>(0.86,1.41) | 0.58<br>(0.39,0.88) | 0.61<br>(0.4,0.93) | 0.67<br>(0.43,1.03) |
| **CSSS support variables** |  |  |  |  |  |
| Effect per additional support session attended |  |  | 2.37<br>(2.16,2.6) | 2.38<br>(2.17,2.62) | 2.46<br>(2.23,2.72) |
| NRT items per session |  |  |  |  |  |
| >1 item (vs. ≤1 item) |  |  | 1.69<br>(1.15,2.48) | 1.68<br>(1.14,2.46) | 1.71<br>(1.16,2.52) |
| **Type of contact with the hospital-based service (vs. Specialist assessment in-person whilst an inpatient)** |  |  |  |  |  |
| Post-discharge specialist assessment or Very Brief Advice |  |  |  | 0.90<br>(0.42,1.96) | 0.78<br>(0.35,1.73) |
| Outpatient specialist assessment or Very Brief Advice |  |  |  | 1.64<br>(0.99,2.72) | 1.62<br>(0.97,2.7) |
| Unknown |  |  |  | 0.94<br>(0.63,1.4) | 0.89<br>(0.6,1.34) |
| **Health variables** |  |  |  |  |  |
| Specific health conditions (vs. no health conditions) |  |  |  |  |  |
| Chronic respiratory condition |  |  |  |  | 0.96<br>(0.61,1.53) |
| Mental ill health |  |  |  |  | 0.87<br>(0.49,1.54) |
| Cardiovascular disease |  |  |  |  | 0.96<br>(0.59,1.56) |
| Diabetes |  |  |  |  | 0.84<br>(0.48,1.49) |
| Cancer |  |  |  |  | 2.26<br>(1.18,4.33) |
| Number of health conditions (vs. no health conditions) |  |  |  |  |  |
| 1–2 |  |  |  |  | 0.81<br>(0.46,1.42) |
| 3+ |  |  |  |  | 0.5<br>(0.2,1.26) |

<sup>1</sup> Missing values imputed; estimates generated from 19 imputed data sets, each with $N = 1,326$ .

#### Model-1: demographic and socio-economic variables

The odds of achieving a 4-week quit increased with age and were lower for people who were eligible for free NHS prescriptions (Table 3). The odds of quitting by someone with free NHS prescriptions were 28% lower than without.

#### Model-2: Model-1 with nicotine dependence score

Adding nicotine dependence to Model-1 did not significantly improve model fit (Table 2).

#### Model-3: Model-2 with CSSS support variables

Adding the number of CSSS support sessions and the use of pharmacotherapy resulted in large improvements in model fit (Table 2), also associated with increased odds of quitting success (Table 3). On average, individuals in our analysis sample attended 6.3 CSSS support sessions (range: 1 to 26). While most people tended to have ≤1 NRT items given per session, 31.0% were on average given more than one type of NRT per session, indicating more intensive pharmacotherapy (see Section 2.6 for the definition of this index). Furthermore, accounting for CSSS support variation removed the statistically significant association between older age and higher odds of quitting. At the same time, the association between having free NHS prescriptions and lower odds of quitting became stronger. Higher nicotine dependence also became significantly associated with lower odds of quitting (Table 3). These changes in association happened because individuals with these characteristics received more CSSS support. Consequently, when we accounted for this extra support, the odds of quitting associated with these characteristics decreased.

#### Model-4: Model-3 with hospital contact type

Adding hospital contact type did not produce a statistically significant improvement in model fit (Table 3). There was a trend for people who arrived at CSSS after a hospital outpatient contact to have relatively higher odds of quitting, but this was not statistically significant (Table 3). Supplementary Table S13 gives the observed percentages of quits according to hospital-based contact type and health condition.

#### Model-5: Model-4 with co-morbid health conditions

Adding information on health conditions significantly improved model fit (Table 2). In our analysis sample, 71.9% of people had at least one health condition, and 25.4% of people had three or more health conditions. The five most common conditions were chronic respiratory conditions (37.6%), mental ill health (26.3%), cardiovascular disease (25.2%), diabetes (12.4%), and cancer (9.3%). Compared to having no health conditions, the model estimated that having cancer significantly increased the odds of quitting by a factor of 2.26 (Table 3). None of the associations between quitting and the other health conditions reached statistical significance. There was a trend towards having a greater number of co-morbid health conditions decreasing the odds of quitting, but not with statistical significance (Table 3).

## 4. DISCUSSION

This study investigated the numbers of people identified by the QUIT hospital-based tobacco dependence treatment service who were then referred to a CSSS, and what patient- and service-related factors were associated with a successful quit attempt. After the initial phase-in of hospital-based services, hospital-referred people accounted for 26% of all referrals to the CSSS. Of the hospital-referred people who then made a CSSS supported quit attempt, 61% achieved a 4-week quit. This quitting rate was slightly below the local average for people quitting with CSSS support, but similar to the national average. Extrapolating these results based on the estimated relapse rates over time for people using NRT in clinical trials,^36^ this 4-week quit rate equates to an expected 6-month quit rate of 33%. This quit rate is similar to the quit rates of ∼35% among people who received the Ottawa style intervention, in which support was given by phone for 6-months post-discharge.^16,17^ Compared to the general cohort of people quitting with CSSS support, those transferred from QUIT hospital tobacco teams to CSSS had a higher representation of older people, lower socio-economic status, and long-term health conditions. Thus, the hospital-based service is likely to be facilitating a greater reach of CSSS to priority population subgroups.

### 4.1. Implications for service improvement

#### Who might require additional support to quit?

While research has identified factors influencing quitting among smokers in general^37^ and those receiving CSSS support,^38,39^ there is limited evidence on how people’s health and hospital contact types are associated with quitting outcomes.^30^ Previous studies have suggested lower odds of quitting success for patients with higher cardiovascular risk,^25^ more co-morbidities,^40^ and certain mental health histories.^41–43^ People referred to CSSS from hospital with poorer health status (e.g. more co-morbid conditions) could have lower capability (e.g. mental strength) to quit, although for others the hospital contact might be a catalyst that supports quitting (e.g. due to a cancer diagnosis).^44^ Our findings showed that people who were eligible for free NHS prescriptions consistently had lower odds of quitting, which matched previous findings from English CSSSs.^39^ Eligibility for free prescriptions depends on a range of health and socio-economic factors, which potentially acts as a proxy for others factors associated with lower odds of quitting (e.g. lower socio-economic status). In addition, whilst ethnicity in our analysis sample was almost entirely “White”, a recent analysis from another English hospital-based tobacco dependence treatment service found that people whose ethnicity was classified as “Mixed/Asian/Other” had a lower chance of quitting successfully.^17^ Given such patient characteristics are associated with lower odds of quitting, additional research and then service tailoring is needed to better understand why, and then support people with these characteristics to quit successfully.

#### The importance of flexible and personalised smoking cessation support

In keeping with the challenges faced by other acute hospitals,^17,45,46^ the QUIT service has experienced challenges with high levels of patient drop-out after leaving hospital.^25^ The findings reveal that out of every 1,000 referrals, the CSSS were unable to contact 280 people, 195 declined support after being contacted, and 111 registered with the service but did not attempt to quit, resulting in a total dropout of 586 people, or 59%. It is important that hospital-based services do what they can to maximise patient motivation to quit, and in doing so increase the likelihood that patients will go on to engage with support to stop smoking outside of the hospital environment. Having the right kind of interaction to prompt someone to think about their smoking is therefore an important part of hospital-based tobacco dependence treatment.^44^ Potentially more challenging is understanding the barriers to making a quit attempt and accessing the right kind of support to stop smoking for people recently discharged from hospital, who might be acutely unwell.

For example, cancer patients can encounter a range of barriers when seeking support to quit smoking, even though they are potentially more motivated.^47^ We found that people with cancer receiving CSSS support had relatively higher odds of quitting, suggesting the current pathway is potentially working well for this patient population. This aligns with recent findings that receiving hospital treatment for a smoking-related disease, which included cancer, increased the probability of not smoking when assessed 6-months after discharge.^17^ These findings suggest the benefits of hospital tobacco teams facilitating referral to CSSSs for people whose hospital contact is potentially smoking-related (e.g. cancer diagnosis/care), as this could be acting as a catalyst that increases their motivation to quit. These benefits of facilitating referral to CSSSs should also apply to stop smoking support for individuals who smoke and attend cancer screening.^48^ In the national English service specification, hospital-based tobacco teams should make personalised plans for patients’ ongoing support to stop smoking prior to their discharge, with follow-up calls at 7–14 days and 28-days after discharge.^13^ The discharge plan and follow-up calls provide an opportunity to highlight the flexibility and choice in support options, which could help facilitate engagement.^24^ They also provide an opportunity to assess whether a patient is ready to be referred to CSSS with a view to making a quit attempt. If a patient is referred, CSSS might also benefit from enhanced information sharing, e.g. about health status and the integration of smoking into care plans. This information would help CSSSs to be able to continue personalised care from hospital, and to tailor their approach to hospital-referred patients in an effort to decrease the initial dropout. Such initiatives to improve the personalisation of care could also help to improve quitting outcomes for other groups supported by hospital-based tobacco teams, such as children, parents and carers of children, and NHS staff who smoke.

### 4.2. Strengths and limitations of the study

Our study’s strengths include its systematic approach to analysis development and conduct using real-world CSSS data. Our statistical analysis plan development was supported by a literature review to identify factors that might be associated with quitting outcomes.^30^ Further details of population eligibility criteria and the covariates of interest were developed in the interim QUIT service evaluation report.^25^ The evaluation team refined the analysis plan through regular discussions with the QUIT service team, including the programme director and manager, through the stages of evaluation planning, data collection, and data curation. These discussions addressed various challenges, including constraints on data collection within service settings, navigating information governance, data sharing and completeness, and understanding the collected data fields. The QUIT service and CSSS teams aided with interpreting the study findings, to provide service-context to our quantitative analyses.

There were four main study limitations, which primarily concern the study and analysis sample. First, we could not investigate quitting outcomes beyond whether someone achieved a 4-week quit, although achieving a 4-week quit is predictive of longer-term quitting success.^36,49^ Second, the size of the data sample was limited because logistical constraints imposed by CSSS commissioning arrangements meant that the study was only able to use data from one of two CSSSs receiving patients from the acute hospitals implementing the QUIT service. This meant that CSSS activity resulting from referrals from The Rotherham Hospital NHS Foundation Trust could not be included in this study. Third, due to data sharing restrictions, it was not possible to use individual-level data to compare the characteristics and quitting outcomes of hospital-referred people to other people quitting with support from the CSSS. To overcome this, we used aggregate summary statistics from the CSSS that provided data for this study and the national CSSS reporting data as the comparator, although the national reporting data has its own data quality limitations and may not be a wholly fair comparator.^29^ Fourth, although we intended to, it was not possible for us to link a sufficiently large sample of individuals between the hospital-based records and the CSSS records. This meant that for our analyses it was not possible to use the information on smoking status and treatment throughout a patient’s stay in hospital, as recommended in the standard evaluation framework for these services (see also Section 4.3).^50^ See the Supplementary Information for a discussion of implications for service monitoring and evaluation.

## 4. CONCLUSION

People referred to CSSS from hospital made up around 26% of all CSSS referrals and may not have attempted to quit before their hospital contact. As such, identifying smokers during a hospital-based contact then offering ongoing smoking cessation support is likely to be increasing the number of people quitting smoking.

However, there was a high drop-out rate between hospital referrals and CSSS. Some patients could not be contacted (28%), and of those who were contacted many declined support or did not then go on to make a quit attempt even after accepting support (43%).

### Actions to improve transfer of care

#### Hospital

- While motivation to quit is assessed pre-referral and patients are followed-up with post-discharge phone calls from the hospital-based team to confirm the wish to be referred, some patients may experience barriers to quitting smoking after their hospital stay. Hospitals should explore ways to maximise patient motivation to quit by understanding the potential barriers to quitting before a referral to CSSS.

#### CSSS

- Hospital-referred patients may have different motivations for quitting than the general CSSS population. CSSS should consider tailoring their initial approach to acknowledge these motivations and potential barriers related to recent hospitalisation.
- Improved information sharing from hospitals could help to personalise CSSS support. Sharing details about a patient’s medical history, social situation, and initial motivations for quitting could help CSSS in tailoring their approach and identifying those who might require additional support.

#### Implications for quitting success

The study found that hospital-referred patients who engaged with CSSS achieved quit rates similar to the general CSSS population. However, quit rates were lower among patients with eligibility for free NHS prescriptions, a potential indicator of long-term health conditions or socio-economic disadvantage. This finding suggests a need for additional support for this subgroup within both hospital and community settings.

Overall, collaboration between hospital and community stop smoking services is key. By jointly focusing on optimising the transfer of care and tailoring support to individual needs, both hospital and community stop smoking services can improve patient engagement and ultimately increase quit rates.

## ACKNOWLEDGMENTS

We would like to thank the teams at the NHS South Yorkshire Integrated Care Board and at the NHS hospital trusts who are implementing the QUIT service. In particular, Dr Lisa Wilkins, the QUIT Programme Director and Leanne Sparks, Senior Prevention Programme Manager, NHS South Yorkshire Integrated Care Board. Thanks to the QUIT Oversight Group and Dr Richard Jenkins, Senior Responsible Officer for the QUIT Programme for support and feedback on the evaluation. Community Stop Smoking Service data was provided by the NHS Yorkshire Smokefree Service, delivered by South West Yorkshire Foundation Trust. Data transfer was facilitated by Laura Boast at the NHS Yorkshire Smokefree service and managed by Andy Eames at the South Yorkshire Integrated Care Board, both of whom provided support with understanding the data and interpreting the findings. We acknowledge helpful ongoing discussions with Paul Lambert from Yorkshire Cancer Research. Comments that helped to improve the manuscript were given by John Holmes, John Robins, Debbie Robson and Nick Woodrow.

## OPEN ACCESS STATEMENT

For the purpose of open access, the author has applied a CC BY public copyright licence to any Author Accepted Manuscript version arising from this submission.

## DATA AVAILABILITY STATEMENT

The data underlying this article cannot be shared publicly to protect the privacy of participants. An anonymised version of the data, restricted to include only the variables used for analysis, will be shared on reasonable request to the corresponding author.

## CONTRIBUTORSHIP STATEMENT

Conceptualization (DG, MF), Data curation (DG, RC), Formal Analysis (RC), Funding acquisition (DG, MF, SB, JC), Methodology (DG, MF), Project administration (DG), Supervision (DG), Validation (DG), Visualization (RC), Writing – original draft (DG, RC), Writing – review & editing (DG, RC, MF, SB, JC).

