## Supplementary Information for "Smoking Cessation after Transitioning from Hospital to Community Stop Smoking Services: Insights from Real-world Data Analysis"

### 1 The QUIT service

The QUIT service (https://sybics-quit.co.uk) is a major investment across the South Yorkshire Integrated Care Board (ICB), which aims to systematically treat tobacco addiction within secondary care. In 2021, 16.1% of adults over age 18 in the ICB area were smokers, compared to 13% in England as a whole.[1]

QUIT is a complex service being delivered in partnership with eight NHS hospital trusts, five community stop smoking services/integrated wellbeing services and community pharmacies. The service spans five local authority areas.

The QUIT service has four main strands (for a detailed service specification see https://sybics-quit.co.uk/healthcare-professionals/quit-treatment-pathways)

- QUIT for Patients: treatment of tobacco addiction by clinical teams and specialist Tobacco Treatment Advisers.
- QUIT for Parents: clinical teams will provide very brief advice and refer the parent of children who are receiving care from the trust to the tobacco treatment advisers.
- QUIT for Staff: on site specialist support from tobacco treatment advisers and free medications for NHS Trust staff who would like to quit.
- QUIT for a Smoke Free Hospital: promoting a smoke free environment for staff, patients, and visitors.

The QUIT for Patients strand is composed of:

- Acute trust inpatients
- Mental health trust inpatients
- Children's inpatients
- Secondary care community mental health
- Acute trust: A&E, outpatients and day cases

**Table S1.** Trusts implementing each patient pathway.

<table>
<tr><th>Patient pathway</th><th>Trust</th></tr>
<tr><td>Acute Trust: Inpatients, A&E, Outpatients and Day Cases</td><td>• Doncaster and Bassetlaw Hospitals NHS Foundation Trust (DBTH)<br>• Barnsley Hospitals NHS Foundation Trust (BHNFT)<br>• The Rotherham Hospital NHS Foundation Trust (TRFT)<br>• Sheffield Teaching Hospitals NHS Foundation Trust (STH)</td></tr>
<tr><td>Mental Health Trust Inpatients</td><td rowspan="2">• Rotherham, Doncaster, and South Humber NHS Foundation Trust (RDASH) (Rotherham and Doncaster psychiatric sites and services only)<br>• Sheffield Health and Social Care NHS Foundation Trust (SHSC)<br>• Southwest Yorkshire Partnership NHS Foundation Trust (Barnsley psychiatric sites and services only) (SWYFT)</td></tr>
<tr><td>Secondary Care Community Mental Health</td></tr>
<tr><td>Children and parents/carers</td><td>• Sheffield Children’s Hospital (SCH)<br><br>And children admitted to 4 Acute Trusts:<br>• Doncaster and Bassetlaw Hospitals NHS Foundation Trust<br>• Barnsley Hospitals NHS Foundation Trust<br>• The Rotherham Hospital NHS Foundation Trust<br>• Sheffield Teaching Hospitals NHS Foundation Trust</td></tr>
</table>

**Table S2.** The letters in QUIT stand for the key steps taken by ward staff to start someone's tobacco addiction treatment when they have been admitted to hospital.

| | | |
|---|---|---|
| Q | Ask the **Q**uestion | Ask all hospital patients is they have smoked in the past month. |
| U | **U**nderstand their addiction | Ask all hospital patients to exhale into a Carbon Monoxide breathalyser.<br><br>Ask patients who smoke about how much they smoke to help work out how much Nicotine Replacement Therapy (NRT) to give to those aged 12 and over. |
| I | **I**nform patients | About smoke-free sites & that specialist support is available while they are in hospital. |
| T | Initiate **T**reatment | Offer all smokers NRT on admission & notify the tobacco treatment advisors of all smokers so they can provide specialist support and treatment as soon as possible. |

### 2 Summary statistics from the NHS Yorkshire Smokefree Community Stop Smoking Service

The NHS Yorkshire Smokefree Community Stop Smoking Service (CSSS) provided summary statistics on the numbers of people referred to and engaging with their service, in total and from the QUIT service from July 2021 to March 2023.

**Table S3.** CSSS statistics on patients referred from hospital. These are summary statistics provided by the NHS Yorkshire Smokefree CSSS and are separate to the analysis data sample. The data in this table correspond to the period July 2021 to March 2023. See Table S4 for a monthly breakdown of the trends in referrals, quit dates and 4-week quits.

| | Count | Percentage |
|---|---|---|
| **Number of referrals from the (QUIT) hospital service to CSSS** | 3,223 | |
| **Contacted** | 2,322 | 72.0% |
| **Accepted** | 1,692 | 72.9% |
| **Set quit date** | 1,333 | 78.8% |
| **4 week quit** | 815 | 61.1% |

**Table S4.** Monthly statistics showing total CSSS activity and the percentage of that activity due to referrals from the QUIT service.

| | Total CSSS clients | | | Clients referred from the QUIT service | | | Percentage of clients referred from the QUIT service | | |
|---|---|---|---|---|---|---|---|---|---|
| **Month** | **Referrals** | **Set a quit date** | **Achieved a 4-week quit** | **Referrals** | **Set a quit date** | **Achieved a 4-week quit** | **Referrals** | **Set a quit date** | **Achieved a 4-week quit** |
| **Jul-21** | 667 | 511 | 327 | 56 | 21 | 12 | 8% | 4% | 4% |
| **Aug-21** | 748 | 460 | 290 | 70 | 29 | 19 | 9% | 6% | 7% |
| **Sep-21** | 870 | 427 | 288 | 59 | 13 | 9 | 7% | 3% | 3% |
| **Oct-21** | 841 | 497 | 331 | 114 | 35 | 29 | 14% | 7% | 9% |
| **Nov-21** | 846 | 522 | 347 | 100 | 45 | 30 | 12% | 9% | 9% |
| **Dec-21** | 472 | 329 | 219 | 107 | 36 | 27 | 23% | 11% | 12% |
| **Jan-22** | 975 | 541 | 375 | 131 | 48 | 24 | 13% | 9% | 6% |
| **Feb-22** | 728 | 465 | 331 | 137 | 63 | 38 | 19% | 14% | 11% |
| **Mar-22** | 807 | 420 | 310 | 154 | 73 | 41 | 19% | 17% | 13% |
| **Apr-22** | 670 | 418 | 303 | 181 | 71 | 47 | 27% | 17% | 16% |
| **May-22** | 778 | 464 | 292 | 208 | 92 | 58 | 27% | 20% | 20% |
| **Jun-22** | 718 | 398 | 246 | 180 | 65 | 26 | 25% | 16% | 11% |
| **Jul-22** | 722 | 411 | 275 | 193 | 72 | 36 | 27% | 18% | 13% |
| **Aug-22** | 769 | 412 | 285 | 194 | 82 | 43 | 25% | 20% | 15% |
| **Sep-22** | 718 | 369 | 272 | 178 | 70 | 43 | 25% | 19% | 16% |
| **Oct-22** | 727 | 440 | 300 | 166 | 69 | 47 | 23% | 16% | 16% |
| **Nov-22** | 693 | 427 | 274 | 200 | 88 | 52 | 29% | 21% | 19% |
| **Dec-22** | 488 | 295 | 198 | 170 | 78 | 53 | 35% | 26% | 27% |
| **Jan-23** | 961 | 508 | 341 | 228 | 96 | 60 | 24% | 19% | 18% |
| **Feb-23** | 799 | 437 | 297 | 198 | 87 | 58 | 25% | 20% | 20% |
| **Mar-23** | 896 | 550 | 352 | 199 | 100 | 63 | 22% | 18% | 18% |
| **Total** | 15,893 | 9,301 | 6,253 | 3,223 | 1,333 | 815 | 20% | 14% | 13% |

#### 3 National CSSS reporting data

Collected by NHS England (previously NHS Digital) from local government commissioned services every 3-months, the data includes the number of quit attempts made and that led to 4-week quits (self-reported and CO verified), individual characteristics (e.g., smoking-related details), and the type of support received.[2] Local profiles were obtained by selecting only the localities covered by the CSSS in this study: Barnsley (identifier E08000016), Doncaster (E08000017), and Sheffield (E08000019), which contained a mix of data from the NHS Yorkshire Smokefree CSSS and other local services, e.g., specialist maternity stop smoking services. However, since 2014, activity reporting by CSSSs to NHS England has been voluntary with no data collection standards; for 2022 to 2023, 15 of 151 local areas did not submit data.[3]

**Table S5. Quit attempts.** Age, sex and socio-economic profile of the number of quit dates set. Comparison between the analysis sample of quit attempts (1st July 2021 to 31st March 2023) and national and local samples (1st April 2022 to 31st March 2023).

| | **Analysis sample** | | **National** | | **Local** [3] | |
|---|---|---|---|---|---|---|
| | **Number of observations** [2] | **Percentage of total sample** | **Number of observations** | **Percentage of total sample** | **Number of observations** | **Percentage of total sample** |
| Total | 1,326 | | 176,566 | | 5,083 | |
| Male | | | | | | |
| Under 18 [1] | - | - | 484 | 0.3% | 15 | 0.3% |
| 18–34 | 62 | 4.7% | 11,640 | 6.6% | 346 | 6.8% |
| 35–44 | 102 | 7.7% | 15,120 | 8.6% | 391 | 7.7% |
| 45–59 | 273 | 20.6% | 28,761 | 16.3% | 832 | 16.4% |
| 60 and over | 254 | 19.2% | 24,045 | 13.6% | 709 | 13.9% |
| Female | | | | | | |
| Under 18 | - | - | 571 | 0.3% | 21 | 0.4% |
| 18–34 | 50 | 3.8% | 23,445 | 13.3% | 585 | 11.5% |
| 35–44 | 88 | 6.6% | 17,626 | 10.0% | 452 | 8.9% |
| 45–59 | 231 | 17.4% | 29,705 | 16.8% | 868 | 17.1% |
| 60 and over | 266 | 20.1% | 25,169 | 14.3% | 864 | 17.0% |
| Occupation | | | | | | |
| Routine and manual occupations | 411 | 31.0% | 42,394 | 24.0% | 1,644 | 32.3% |
| Retired | 321 | 24.2% | 25,642 | 14.5% | 908 | 17.9% |
| Sick/disabled and unable to work | 296 | 22.3% | 22,233 | 12.6% | 839 | 16.5% |
| Never worked or unemployed for over 1 year | 196 | 14.8% | 26,376 | 14.9% | 851 | 16.7% |
| Other | 102 | 7.7% | 59,921 | 33.9% | 841 | 16.5% |
| Eligible for free NHS prescriptions | | | | | | |
| Yes | 1,076 | 81.1% | 109,015 | 61.7% | 3,034 | 59.7% |

[1] The analysis sample was restricted to individuals 18 years and over, excluding 2 individuals.

[2] The number of observations does not necessarily equal the number of individuals because individuals could make more than one quit attempt.

[3] Local profiles were obtained by selecting only the localities covered by the CSSS in this study: Barnsley (identifier E08000016), Doncaster (E08000017) and Sheffield (E08000019).

**Table S6. Quit success.** Age, sex and socio-economic profile of the number of 4-week quits. Comparison between the analysis sample (1st July 2021 to 31st March 2023) and national and local samples (1st April 2022 to 31st March 2023).

| | **Analysis sample** | | **National** | | **Local [3]** | |
|---|---|---|---|---|---|---|
| | **Number of 4-week quits [1]** | **Percentage of quit attempts** | **Number of 4-week quits** | **Percentage of quit attempts** | **Number of 4-week quits** | **Percentage of quit attempts** |
| Total | 813 | 61.3% | 109,356 | 61.9% | 3,540 | 69.6% |
| Male | | | | | | |
| Under 18 | - | - | 269 | 55.6% | 11 | 73.3% |
| 18–34 | 29 | 46.8% | 6,943 | 59.6% | 222 | 64.2% |
| 35–44 | 57 | 55.9% | 9,379 | 62.0% | 254 | 65.0% |
| 45–59 | 167 | 61.2% | 18,436 | 64.1% | 566 | 68.0% |
| 60 and over | 177 | 69.7% | 16,176 | 67.3% | 529 | 74.6% |
| Female | | | | | | |
| Under 18 | - | - | 353 | 61.8% | 23 | 109.5% [3] |
| 18–34 | 24 | 48.0% | 13,768 | 58.7% | 509 | 87.0% |
| 35–44 | 53 | 60.2% | 10,418 | 59.1% | 309 | 68.4% |
| 45–59 | 137 | 59.3% | 17,925 | 60.3% | 532 | 61.3% |
| 60 and over | 169 | 63.5% | 15,689 | 62.3% | 585 | 67.7% |
| Occupation [2] | | | | | | |
| Routine and manual occupations | 254 | 61.8% | 24,114 | 56.9% | 1,084 | 65.9% |
| Retired | 213 | 66.4% | 14,701 | 57.3% | 628 | 69.2% |
| Sick/disabled and unable to work | 171 | 57.8% | 11,753 | 52.9% | 514 | 61.3% |
| Never worked or unemployed for over 1 year | 104 | 53.1% | 12,716 | 48.2% | 497 | 58.4% |
| Other | 71 | 69.6% | 32,116 | 53.6% | 539 | 64.1% |
| Eligible for free NHS prescriptions | | | | | | |
| Yes | 649 | 60.3% | 58,906 | 54.0% | 1,889 | 62.3% |

[1] The sum of self-reported and carbon monoxide verified 4-week quits.

[2] Only 87.2% (95,400 / 109,356) of 4-week quits in the national data had data on occupation; 4-week quits without occupation are not reported.

[3] A more than 100% quit success is likely to be due to data recording error, which is made more obvious due to the small number of quit attempts.

### 4 Further details of explanatory variables for regression analysis

**Table S7.** Socio-economic classifications used in the stop smoking service data. Categories were merged due to limited sample size.

| Variable definitions in the service monitoring data[1] | Analysis category |
|---|---|
| Routine and manual (self-employed should not be included in this category). Examples include: electrician, fitter, gardener, inspector, plumber, printer, train driver, tool maker, bar staff, caretaker, catering assistant, cleaner, farm worker, HGV driver, labourer, machine operative, messenger, packer, porter, postal worker, receptionist, sales assistant, security guard, sewing machinist, van driver, waiter/waitress. | Routine and manual occupations |
| Retired | Retired |
| Sick or disabled and unable to work | Sick or disabled and unable to work |
| Never worked or unemployed for over one year. If unemployed for less than one year, last known occupation should be used for classification. | Never worked or unemployed for over 1 year |
| Full-time Student | Other |
| Home carer i.e. looking after children, family or home | Other |
| Managerial/ professional. Examples include: accountant, artist, civil/mechanical engineer, medical practitioner, musician, nurse, police officer (sergeant or above), physiotherapist, scientist, social worker, software engineer, solicitor, teacher, welfare officer. Those usually responsible for planning, organising and co-ordinating work for finance | Other |
| Intermediate. Examples include: call centre agent, clerical worker, nursery auxiliary, office clerk, secretary | Other |
| Prisoner | Other |
| Other (not listed) | Other |

[1] Available from: https://www.datadictionary.nhs.uk/data_elements/socio-economic_classification_code__stop_smoking_.html

**Table S8.** Names of the medical conditions available in the data and their grouping to form the categories of disease that were used for analysis.

| **Medical condition** | **Category** |
|---|---|
| Angina | Cardiovascular disease |
| Heart attack | Cardiovascular disease |
| Heart disease / stroke | Cardiovascular disease |
| Diabetes | Diabetes |
| Depression | Mental ill health |
| Anxiety | Mental ill health |
| Severe mental illness | Mental ill health |
| Other long-term mental health condition | Mental ill health |
| Cancer | Cancer |
| Asthma | Chronic respiratory conditions |
| Chronic obstructive pulmonary disease | Chronic respiratory conditions |
| Epilepsy | Not investigated individually due to insufficient data but used in the description of multi-morbidity. |
| Renal disease | Not investigated individually due to insufficient data but used in the description of multi-morbidity. |
| Thyroid - overactive | Not investigated individually due to insufficient data but used in the description of multi-morbidity. |
| Post & pre-operative | Not investigated individually due to insufficient data but used in the description of multi-morbidity. |
| Any other long-term condition | Not investigated individually due to insufficient data but used in the description of multi-morbidity. |

**Table S9.** Details for why some variables that were intended to be included in the analysis were not included.

| **Variable intended for inclusion** [4] | **Reason not included** |
|---|---|
| Index of Multiple Deprivation (IMD) (a small-area geographic indicator of socio-economic conditions) [5] | Postcode information only provided to the level of sector of residence. Full postcode needed to link to IMD. |
| Ethnicity | Available as a 17-level classification.<br>Not usable because majority white or unknown.<br>Sample: 85% (1,425 / 1,677) white, 11% (186 / 1,677) missing. |
| Maternity status | Available as pregnant yes/no. Not usable because majority no. |
| Lives with other smokers | Available as yes/no. Not usable because 41% missing. |
| Number of previous attempts to stop smoking | Not available |
| Type of inpatient admission (elective/emergency) | CSSS data could not be linked to inpatient hospital records. |
| Speciality of the medical consultant caring for the patient | CSSS data could not be linked to inpatient hospital records. |
| Primary diagnosis (the diagnosis assigned on admission to hospital) | CSSS data could not be linked to inpatient hospital records. |
| Whether on medication whose metabolism was affected by smoking | Available as yes/no. Not usable because 94% missing. |
| Whether attained a continuous four-week period of abstinence while hospitalised | Available as yes/no. Not usable because 97% missing. |

### 5 Flows of patients through the hospital based (QUIT) service and CSSS

To describe the flow of patients through the main inpatient tobacco dependence treatment pathway during the study period, we used summary statistics from the monthly Key Performance Indicators for the QUIT service, compiled by the South Yorkshire Integrated Care Board for monitoring purposes. Due to delays in setting up these monitoring statistics, data from all three hospitals in this study were only available from November 2022 to March 2023. These data include the number of hospital admissions, patients identified as current smokers, contacts with hospital-based tobacco treatment advisors, locally resident contacts, and resulting CSSS referrals.

**Table S10**. Main hospital inpatient pathway for the three acute hospitals that referred patients to the CSSS in this study. Data from 1st November 2022 to 31st March 2023. Data is limited to inpatients aged 16 and over with a length of stay >1 night. These figures are an excerpt from the monthly Key Performance Indicator data collated by the South Yorkshire Integrated Care Board for service monitoring purposes.

| | **Barnsley Hospital NHS Foundation Trust** | | **Doncaster and Bassetlaw Hospitals NHS Foundation Trust** | | **Sheffield Teaching Hospitals NHS Foundation Trust** | | **Total** | |
|---|---|---|---|---|---|---|---|---|
| | **Number** | **%** | **Number** | **%** | **Number** | **%** | **Number** | **%** |
| Number of admissions | 8,974 | | 11,084 | | 25,546 | | 45,604 | |
| Number identified as a smoker [1] | 868 | 9.7% | 1,638 | 14.8% | 1,910 | 7.5% | 4,416 | 9.7% |
| Number of hospital-based tobacco treatment advisor contacts [2] | 337 | 38.8% | 339 | 20.7% | 1,204 | 63.0% | 1,880 | 42.6% |
| Number of hospital-based tobacco treatment advisor contacts, limited to the South Yorkshire and Bassetlaw Clinical Commissioning Group area [3] | 329 | 97.6% | 222 | 65.5% | 1,159 | 96.3% | 1,710 | 91.0% |
| Number referred to CSSS [3] | 130 | 39.5% | 83 | 37.4% | 356 | 30.7% | 569 | 33.3% |

[1] This refers to smokers identified by either nursing staff or a hospital-based tobacco dependence treatment advisor at any point during their admission.

[2] This could be a specialist assessment either in-person whilst an inpatient or a post-discharge phone call.

[3] This refers to people who were registered with a General Practitioner within the South Yorkshire and Bassetlaw Clinical Commissioning Group (CCG) area. Whilst CCGs as an organisational entity have since been replaced by Integrated Care Boards, whether someone was registered in the local CCG area is a useful indicator that they are likely to attend the local CSSS, as opposed to a CSSS in another part of the country.

**Table S11**. Estimated flows of patients through the joint hospital and community stop smoking service pathway. Flows through the hospital-based pathway are taken from Table S10 and flows through the community pathway are taken from Table S3.

| | **Details** | **Percentage** | **Standardised number** |
|---|---|---|---|
| Admissions | Patients aged 16+ years with a length of stay in hospital of at least one day. | | 100,000 |
| Patients identified as currently smoking tobacco | Identified as someone who smokes by nursing staff on admission or by a hospital-based tobacco dependence treatment advisor at any point during their admission. | 9.7% | 9,700 |
| Tobacco treatment advisor contacts | Patients who are seen by a hospital-based tobacco dependence treatment advisor while an inpatient or phoned within 5 days post-discharge who have a specialist assessment by the hospital-based advisor. | 42.6% | 4,130 |
| Contacts from the local area | Patients who are resident within the South Yorkshire and Bassetlaw Clinical Commissioning Group area. | 91.0% | 3,760 |
| Referred to CSSS | Locally resident patients who after contact with the hospital-based tobacco dependence treatment advisor had their care for tobacco addiction transferred onto a CSSS after discharge. | 33.3% | 1,250 |
| Contactable by the CSSS | The CSSS attempts to contact all patients referred from the hospital-based service, but not all can be contacted. | 72.0% | 900 |
| Registered with the CSSS | If the CSSS can contact the patient, and they accept the offer of support, then they are registered as a client with the CSSS. | 72.9% | 660 |
| Set a quit date | Clients registered with the CSSS who go on to make a supported quit attempt. | 78.8% | 520 |
| 4-week quit | Clients who make a supported quit attempt who are recorded as having achieved a 4-week quit. | 61.1% | 320 |

#### 6 Descriptive statistics of the analysis sample

The full data sample comprised 1,677 individuals referred from the QUIT service, who accepted the offer of support and were registered with the CSSS. From this sample, 1,641 individuals (98%) were retained, after excluding those younger than age 18 (n=2), parents/carers of children admitted to hospital (n=24) or hospital-based NHS staff (n=10). For analysis, we further restricted this sample to 1,332 individuals (81%) who received at least one support session from a CSSS advisor and set a start date for their quit attempt. We also excluded 6 individuals who had missing data on their occupation, which left an analysis sample of 1,326. Supplementary Figure S1 shows monthly counts of all individuals registered with the CSSS, of individuals in the analysis sample, and of individuals who achieved a 4-week quit.

**Figure S1:** Description of the CSSS data sample—Monthly counts of registered, in-analysis, and 4 week quits.

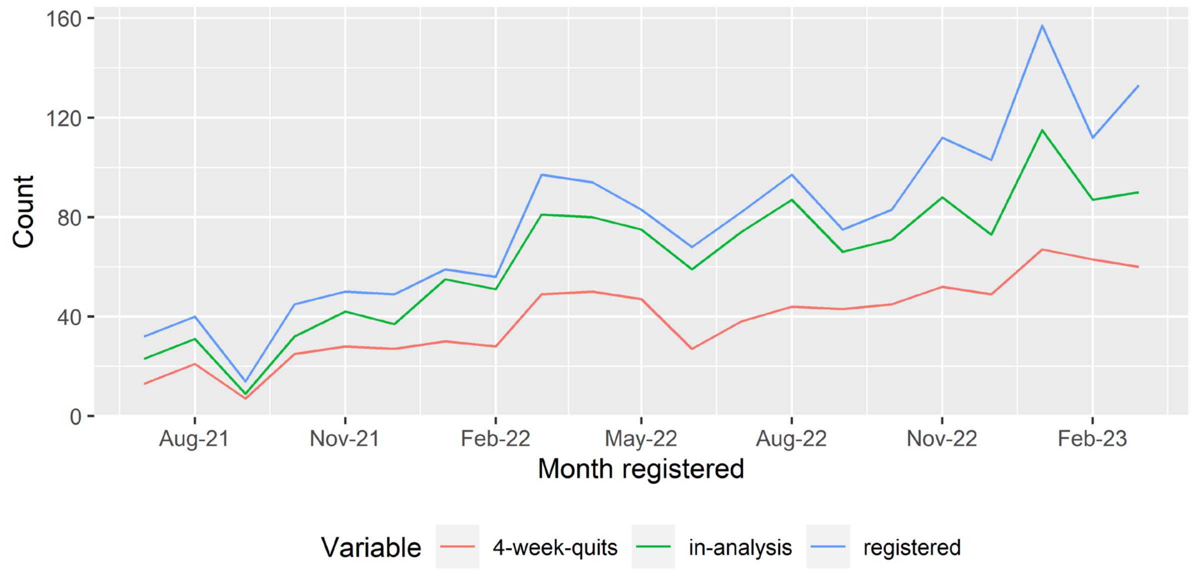

### 7 Supplementary results of statistical analysis

**Table S12.** Sensitivity analysis of the parameter estimates in the base case version of Model 5 (1), to the effects of only including people with complete information on the Fagerström score of nicotine dependence (2), and omitting people lost to follow-up (3).

| | Base case (1) | Complete Fagerström score (2) | Complete follow-up (3) |
|---|---|---|---|
| Constant | 6.72 | 0.01 | 0.04 |
| | (2.91,15.52) | (0,0.04) | (0.01,0.09) |
| **Demographic and socio-economic variables** | | | |
| Male (vs. Female) | 0.78 | 0.75 | 0.84 |
| | (0.55,1.11) | (0.5,1.13) | (0.58,1.21) |
| Age group, reference 18–34 | | | |
| 35–49 | 1.78 | 1.45 | 1.77 |
| | (0.85,3.73) | (0.63,3.39) | (0.8,3.94) |
| 50–59 | 1.3 | 0.94 | 1.23 |
| | (0.67,2.51) | (0.44,1.99) | (0.61,2.49) |
| 60+ | 1.82 | 1.17 | 1.53 |
| | (0.87,3.79) | (0.5,2.74) | (0.7,3.34) |
| Occupation (vs. Routine and manual) | | | |
| Retired | 0.79 | 0.86 | 0.71 |
| | (0.46,1.36) | (0.46,1.59) | (0.4,1.24) |
| Sick/disabled and unable to work | 1.19 | 1.08 | 1.33 |
| | (0.58,2.43) | (0.46,2.5) | (0.62,2.86) |
| Never worked/long term unemployed | 0.79 | 0.77 | 0.8 |
| | (0.45,1.39) | (0.41,1.46) | (0.45,1.44) |
| Other | 0.77 | 0.88 | 0.82 |
| | (0.46,1.31) | (0.48,1.62) | (0.47,1.43) |
| Eligible for free NHS prescriptions (vs. not) | 0.55 | 0.6 | 0.58 |
| | (0.33,0.92) | (0.33,1.08) | (0.34,0.98) |
| Fagerström score for nicotine dependence | | | |
| 5+ (vs. 1–4) | 0.67 | 0.69 | 0.63 |
| | (0.43,1.03) | (0.45,1.06) | (0.4,0.98) |
| **CSSS support variables** | | | |
| Effect per additional support session attended | 2.46 | 2.8 | 2.37 |
| | (2.23,2.72) | (2.49,3.2) | (2.14,2.62) |
| NRT items per session | | | 1.71 |
| >1 item (vs. ≤1 item) | (1.16,2.52) | (1.17,2.92) | (1.27,2.95) |
| | 2.46 | 2.8 | 2.37 |
| **Type of contact with the hospital-based service** (vs. Specialist assessment in-person whilst an inpatient) | | | |
| QUIT type, reference inpatient; full assessment | | | |
| Post-discharge specialist assessment or Very Brief Advice | 0.78 | 0.69 | 0.72 |
| | (0.35,1.73) | (0.28,1.65) | (0.32,1.62) |
| Outpatient specialist assessment or Very Brief Advice | 1.62 | 1.49 | 1.42 |
| | (0.97,2.7) | (0.83,2.71) | (0.85,2.38) |
| Unknown | 0.89 | 0.75 | 0.95 |
| | (0.6,1.34) | (0.47,1.2) | (0.62,1.45) |
| **Health variables** | | | |
| Specific health conditions (vs. no health conditions) | | | |
| Chronic respiratory condition | 0.96 | 0.9 | 1.03 |
| | (0.61,1.53) | (0.54,1.51) | (0.63,1.66) |
| Mental ill health | 0.87 | 0.75 | 0.93 |
| | (0.49,1.54) | (0.39,1.44) | (0.51,1.7) |
| Cardiovascular disease | 0.96 | 0.89 | 0.87 |
| | (0.59,1.56) | (0.51,1.54) | (0.52,1.44) |
| Diabetes | 0.84 | 0.95 | 0.92 |
| | (0.48,1.49) | (0.5,1.8) | (0.5,1.69) |
| Cancer | 2.26 | 2.13 | 1.97 |
| | (1.18,4.33) | (1.01,4.54) | (1.01,3.85) |
| Number of health conditions (vs. no health conditions) | | | |
| 1–2 | 0.81 | 0.91 | 0.77 |
| | (0.46,1.42) | (0.46,1.77) | (0.42,1.4) |
| 3+ | 0.5 | 0.49 | 0.45 |
| | (0.2,1.26) | (0.17,1.41) | (0.17,1.19) |
| Lost to follow up | not quit | not quit | omitted |
| Fagerström dummy imputed | yes | no | yes |
| N | 1,326 | 1,074 | 1,195 |

Estimates are odds ratios. Figures in parenthesis are 95% confidence intervals.

**Table S13.** The percentages of people who made a quit attempt with CSSS, in the analysis data sample, who achieved a 4-week quit according to nature of contact with the hospital-based (QUIT) service and health status.

| **Type of contact with the hospital-based service** | **% of 4 week quits** (assuming lost to follow-up is not quit) | **% of 4 week quits** (excluding lost to follow-up from the denominator) |
|---|---|---|
| Specialist assessment in-person whilst an inpatient | 57.0% | 65.0% |
| Specialist assessment / Very Brief Advice post-discharge | 50.0% | 57.9% |
| Specialist assessment / Very Brief Advice outpatient | 61.4% | 63.9% |
| Unknown | 66.8% | 73.8% |
| **Health variables** | | |
| Specific health conditions | | |
| Chronic respiratory condition | 60.2% | 65.9% |
| Mental ill health | 62.6% | 65.9% |
| Cardiovascular disease | 66.7% | 70.9% |
| Diabetes | 60.0% | 68.8% |
| Cancer | 69.6% | 73.1% |
| Number of health conditions | | |
| 0 | 55.6% | 64.8% |
| 1–2 | 64.6% | 70.3% |
| 3+ | 60.7% | 66.6% |

### 8 Extended implications

**Implications for service monitoring and evaluation**

***Quitting outcomes***

The findings of this study offer a snapshot of the quits obtained via CSSS in an early period of QUIT service implementation, during which the hospital and CSSS teams were actively implementing improvements, particularly in facilitating the effective transfer of patients to CSSS. Further data collection is important to evaluate the service's efficacy once it is fully embedded in the hospital culture and more issues are addressed. A larger dataset would also provide greater statistical power to detect variations in quitting outcomes based on patient and service characteristics.

Monitoring quitting outcomes presents challenges due to differing levels of engagement with both hospital-based services and CSSS. Of the inpatients who received a specialist assessment for tobacco dependence in hospital during the study period, only a third were recorded in the Key Performance Indicator data for the QUIT service to have been referred on to CSSS, which could have been due to them declining or referrals not being recorded. Quitting outcomes in the remainder, i.e. people who were not referred to a CSSS or who were referred but did not engage with the CSSS, have to be ascertained by the hospital tobacco teams in their 4-week post-discharge follow-up appointments, with quitting outcome data from these follow-ups not available for analysis. In addition, quitting outcomes for outpatients and those receiving only Very Brief Advice in hospital could remain unknown unless they go on to take up support from CSSS, as some did in our data sample. The overall quits achieved by the QUIT service will depend on the quitting outcomes of patients who receive all types of post-discharge support, including from CSSS, community pharmacies, and self-quitting.[6] Furthermore, the family and friends of individuals who received support from the QUIT service might have been motivated to quit, but no related data is available. However, as more people arrive at CSSS from hospital, the more representative CSSS outcomes will become of the hospital-based service as a whole. To prevent people from dropping out of smoking cessation support after leaving the hospital and to keep them motivated throughout the process, the QUIT service is exploring new strategies. One approach is to have hospital-based services book CSSS appointments for patients immediately, rather than relying on the CSSS to contact patients after discharge. Additionally, having in-reach CSSS advisors in the hospital to facilitate transfers is being considered.

***Linking patient hospital and CSSS records***

Being able to track the experiences of individuals across hospital and CSSS records could help to understand how improvements to the hospital-based service affect the experiences of people attending CSSS. The original evaluation plan was to analyse predictors of quitting outcomes using a dataset that linked individual records from the hospital-based service to CSSS records using each patient's unique NHS number. This linkage would have allowed for additional explanatory variables to be drawn from the hospital records, adding to the information from the hospital records that was already transferred to the CSSS on patient referral. However, the number of individuals who could be linked between the two record systems was too limited to provide a sample size large enough for meaningful analysis: out of 1,641 individuals in the CSSS data, only 786 had a unique identifier that could be used for linkage. Allowing for a three-week window between hospital discharge and CSSS registration, only 332 individuals could then be linked between the two record systems. Therefore, the explanatory variables for this study were limited to CSSS data only; Supplementary Table S9 gives details for why some intended variables were not included. Improving this data linkage could be beneficial for future service evaluation.

***Monitoring community stop smoking service workload***

Although there are opportunities to support quit attempts by transferring people from hospitals to CSSSs, CSSSs need sufficient staff to deal with the increased number of clients for the service to work appropriately. To estimate demand from our study's findings, we used summary statistics from the CSSS on the number of hospital-referred clients they supported in relation to their total activity. After an initial phase-in, hospital-referred quit attempts accounted for 19% of all quit attempts supported by CSSS, equating to 81 people who attempted to quit who might not have done so without the QUIT service. However, this calculation does not consider the additional work required from CSSS to contact all hospital-referred patients, of whom only 42% (81/191) went on to make a supported attempt to stop smoking.
